# Psychometric evaluation of the Danish Near-Death Experience Content scale in a sample of out-of-hospital cardiac arrest survivors

**DOI:** 10.1101/2025.11.18.25340402

**Authors:** Tobias Anker Stripp, Niels Christian Hvidt

## Abstract

Some individuals have profound and specific experiences during life-threatening events such as cardiac arrest. Such experiences, often termed near-death experiences (NDE), or recalled experiences of death, have been studied extensively internationally, but research in a Scandinavian context is almost nonexistent. No psychometrically tested questionnaire for capturing NDE phenomena exist in any Scandinavian language. This hinders prevalence, cross-cultural, and clinical care studies of NDEs in a Scandinavian context. The aim of the present study was to evaluate the psychometric properties of the Danish translation of the NDE content scale (NDE-C) among out-of-hospital cardiac arrest (OHCA) survivors. A cross-sectional survey was conducted among OHCA survivors (2016–2020) identified through the Danish Cardiac Arrest Register. Participants reporting memories from their cardiac arrest completed the 20-item Danish NDE-C. Of 2,262 invited survivors, 855 responded (37.8%) with 226 participants who completed the NDE-C. The scale demonstrated excellent internal consistency (Cronbach’s α = 0.92). A confirmatory factor analysis supported a five-factor structure conceptually, but global fit indices were mixed with most indices falling below predefined thresholds for a good fit (CFI = 0.858; TLI = 0.831; RMSEA = 0.093; SRMR = 0.062). Factor loadings and discriminant validity were satisfactory. The Danish NDE-C scale shows strong internal consistency and a preliminary acceptable, although not good, factor structure, indicating potential promise as a tool for assessing NDE content in clinical and research contexts. Further psychometric validation is necessary. This marks an important step in the systematic research of NDE and related spiritually transformative experiences in a Scandinavian context.

## INTRODUCTION

The Near-Death Experience (NDE) represents a profound subjective phenomenon reported by individuals who have approached actual or perceived death. NDEs have been systematically studied for more than 50 years, and historical documents show that multiple cultures around the world have known, described, and used NDEs as epistemic tools for millennia ^1^. NDEs are likely to have been central experiences in the development of cultural and religious ideas of the afterlife throughout human history ^2^.

These reports are often characterized by a consistent pattern of features, including sensations of leaving the body (out-of-body experiences) with perceptions from an external vantage point; traveling through darkness and emerging into light, often by way of a tunnel; witnessing another realm; meeting deceased loved ones; encountering the divine; experiencing a life review; deciding to return or being sent back to the body and earthly life ^3^. In fact, more than 50 narrative themes have been identified ^4^. The experience is usually highly spiritual or religious in nature (e.g., related to transcendence of common earthly immanent experience ^5^). Prospective studies report varying prevalences of NDEs for in-hospital (6.3–39.3%) versus out-of-hospital (18.9–21.2%) cardiac arrest ^6^. Core NDE features appear to be universal, although some content exhibits slight variation, probably because of interpretations through individual cultural and societal lenses ^7^. Further, although primarily reported and researched as positive experiences, scholarly work within the last decades shows that some NDErs also report negative or disturbing content during their experience (reports estimate 14% of NDEs contain ‘negative’ content) ^8^. Negative experiences have been reported to be more prevalent in NDEs occurring during suicide attempts, raising the question of top-down processes of individual context affecting the NDE ^8^ – not unlike what Shushan argues as a feasible explanation for slight cultural differences in NDE content ^7^. It is, however, important to notice that the overall valence can be nuanced, with ‘positive’ narrative arcs embedded in NDEs with ‘negative’ content ^8,9^. Whether these negative experiences are a reflection of the NDE itself or because of events surrounding the NDE, such as misinterpretations of real medical intervention such as cardiopulmonary resuscitation, remains debated ^4,10^. In summary, NDEs hold immediate significance for both clinical medicine and metaphysical thinking at the borders of science and faith. Clinically, NDE experiencers (NDErs) often need someone to talk to ^11^, and NDEs are associated with lasting aftereffects necessitating specialized, informed care for experiencers. Metaphysically, if NDE narratives, e.g. of extrasensory veridical perception are considered valid, they challenge reductionist and materialist theories of perceptive consciousness as dependent on the brain by suggesting the potential for organized non-local subjective experience during periods of impaired cerebral function.

The Greyson Near-Death Experience Scale has, since 1983, been the standard for studying NDEs ^12^. It comprises 16 items across four content dimensions with four items in each: affective, cognitive, paranormal, and transcendent ^12^. Recently, researchers reconfigured the Greyson NDE scale as the 20-item Near-Death Experience Content Scale (NDE-C) in an attempt to improve psychometric properties and to include an item on distressing/negative content ^8,13^. Confirmatory factor analyses for the NDE-C supported a structure with five dimensions that relate strongly to the Greyson scale dimensions with some adjustments: *Beyond the usual* is concerned with paranormal sensations, veridical perceptions, speeded thoughts and ineffability. *Insight* covers life review, encountering a presence, sudden understanding, and precognition. *Border* is the sense of reaching a point-of-no return, leaving the earthly world, and experiences of fear/void/non-existence often interpreted as being “negative” content. *Harmony* taps the affective experience of peacefulness and unity. Finally, *Gateway* is concerned with seeing a bright light and traveling through a gateway such as a door or tunnel.

There is currently no validated psychometric instrument in any Scandinavian language. The Greyson NDE scale has been translated into Norwegian, but never psychometrically tested ^14^. This absence severely limits local prevalence studies, cross-cultural comparative analysis, and implementation of evidence-based support for NDErs in this region.

The objectives of this exploratory cross-sectional survey study among out-of-hospital (OH) cardiac arrest (CA) survivors were to provide an initial psychometric evaluation of the Danish translation of the NDE-C by estimating Cronbach’s α and testing the structural validaty through a confirmatory factor analysis (CFA).

## MATERIALS AND METHODS

This study is based on survivors of out-of-hospital cardiac arrest (OHCA) between 2016 and 2020 (n=2,785) identified through the Danish Cardiac Arrest Register, participating in the EXICODE study (a national survey linked to administrative health register data) ^15^. Survivors were invited via secure digital mail to the EXICODE survey, which included a unique question about having any memories from a life-threatening situation such as their cardiac arrest: “A life-threatening situation or illness (such as an accident, cardiac arrest, or cancer) can cause some people to lose or experience an altered sense of consciousness. Some may subsequently recount memories from the time when they had lost, or experienced an altered state of consciousness, whereas others cannot remember anything. Do you have any memories from such an experience?” ^16^. If responding “yes” or “don’t know”, they were presented with a question panel including the NDE-C. Data were collected between the 1st of November and the 13th of December 2021 with a single reminder. Participants were excluded from the main analyses if they had not responded to the EXICODE question probing memories during their cardiac arrest.

### Measures

The primary outcome was the intensity of near-death experiences, measured using the 20-item NDE-C scale ^13^. The translation of the NDE-C into Danish and its qualitative testing are reported elsewhere ^16^. The scale is a Likert-type instrument with response options “0 - not at all; none”, “1 - slightly”, “2 - moderately”, “3 - strongly; equivalent in degree to any other strong experience”, and “4 - extremely; more than any other time in my life and stronger than 3”. The score is the sum across all items (range 0-80). In the development study, the authors reported an overall Cronbach’s α of 0.85 and suggested a cutoff for having an NDE at ≥27 points ^13^.

Demographic data was drawn from registers (2019). Age and sex were determined from the unique 10-digit personal identity number that all Danish citizens have ^17^. Age was categorised into the following groups: 18-35, 36-45, 46-55, 56-65, 66-75, and ≥75. Sex was considered binary as either male (0) or female (1). Education was measured in years and categorised as 7-11, 12-14, and 15+ years. Income was based on household income and coded in tertiles (low, middle, high) relative to age. Cohabitation was categorised as living with someone (partner) versus living alone. Work status was categorised as either working, receiving benefits (student, public) or pension (private, public). Civil status was classified as married, divorced, or unmarried. Chronic disease burden was operationalised as having none, one, or more than one of eight common chronic diseases (asthma, dementia, chronic obstructive pulmonary disease, arthritis, osteoporosis, schizophrenia, type 1-diabetes, type 2-diabetes).

### Statistical Analyses

Descriptive statistics were presented and tested using Pearson’s chi-squared tests for categorical and t-tests for continuous variables. The psychometric properties of the NDE-C were tested by Cronbach’s α and CFA in a structural equation model framework testing both four- and five-factor models with item-factor relations informed by the original validation article, using only full NDE-C datasets. Goodness-of-fit was assessed by the following measures and associated predefined thresholds for a good fit: root mean squared error of approximation (RMSEA): <0.08; standardized root-mean squared residual (SRMR): <0.08; Tucker-Lewis index (TLI): ≥0.95; comparative fit index (CFI): ≥0.90 ^18,19^. Only register variables had missing values (0-1.8%). Missing categorical variables were imputed to the majority category. Missing values for continuous variables (0-2%) were set to the overall mean level. All statistical analyses were performed using STATA 18 with a 5% significance level.

### Ethics

The project was registered at SDU Research & Innovation Organisation (RIO) (journal number: 10.367), the Danish Regional Scientific Ethics Committee (journal number: 20202000-116), and approved by SDU Research Ethics Committee (REC) (journal number: 20/39546). Participants were informed of the content and purpose of the study before participating. Participation was voluntary based on the information provided.

## RESULTS

In total, 2,262 individuals received the invitation (n=523 did not receive the invitation for different reasons). Of the invited, 855 OHCA survivors responded (response rate: 37.8%), with 738 individuals included in final analyses (n=117 had missing data on the question of memory during life-threatening illness). The mean age was 63.6 years (SD: 10.7), with 80% responders being male. The mean overall score of the NDE-C was 14.7 (SD: 15.3) with the mean score for those with overall scores ≥27 being 40.3 (SD: 12.3). The distribution of responses can be seen in Table 1, which also includes both Danish and English item versions.

**Table 1:**
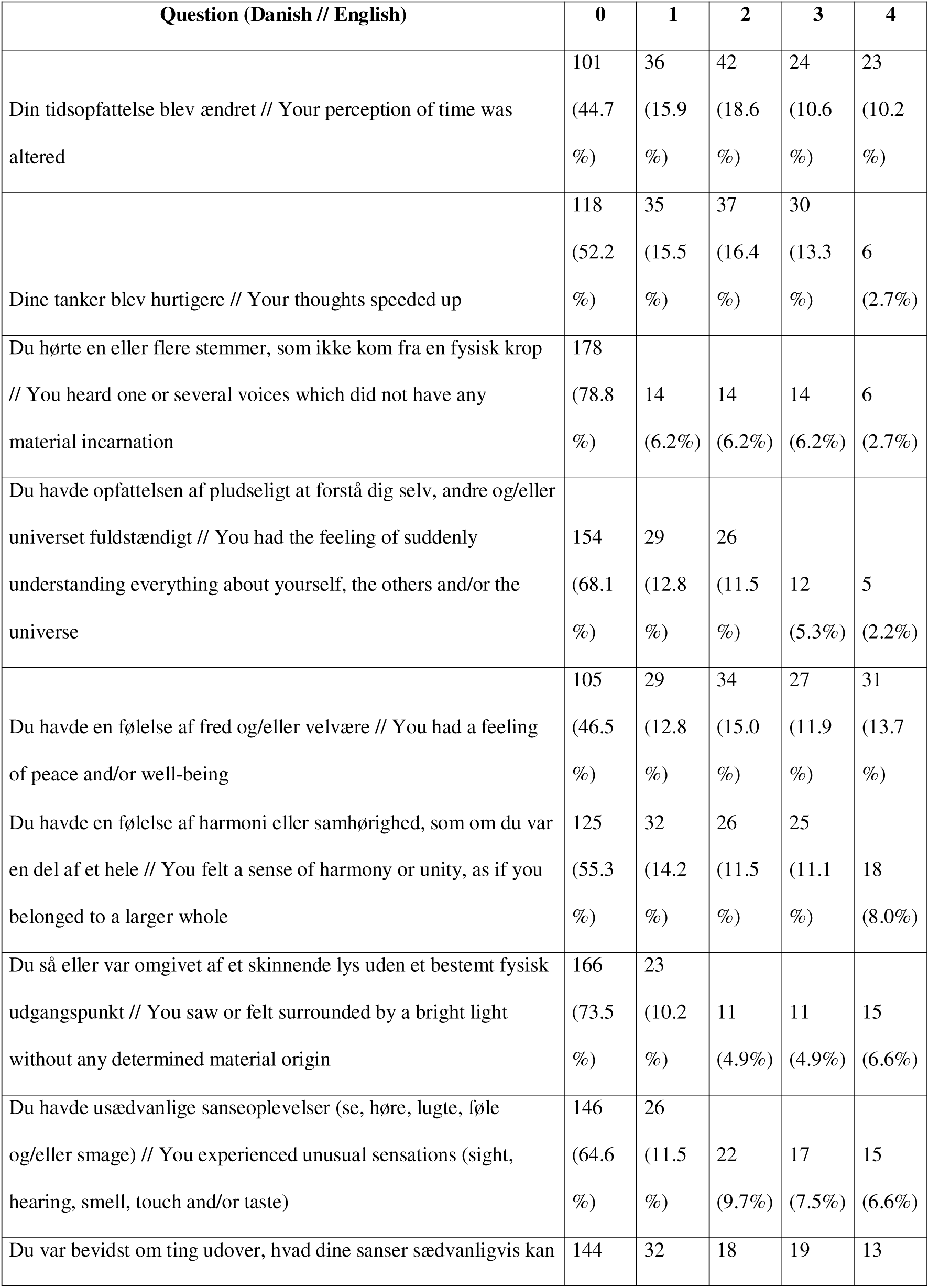
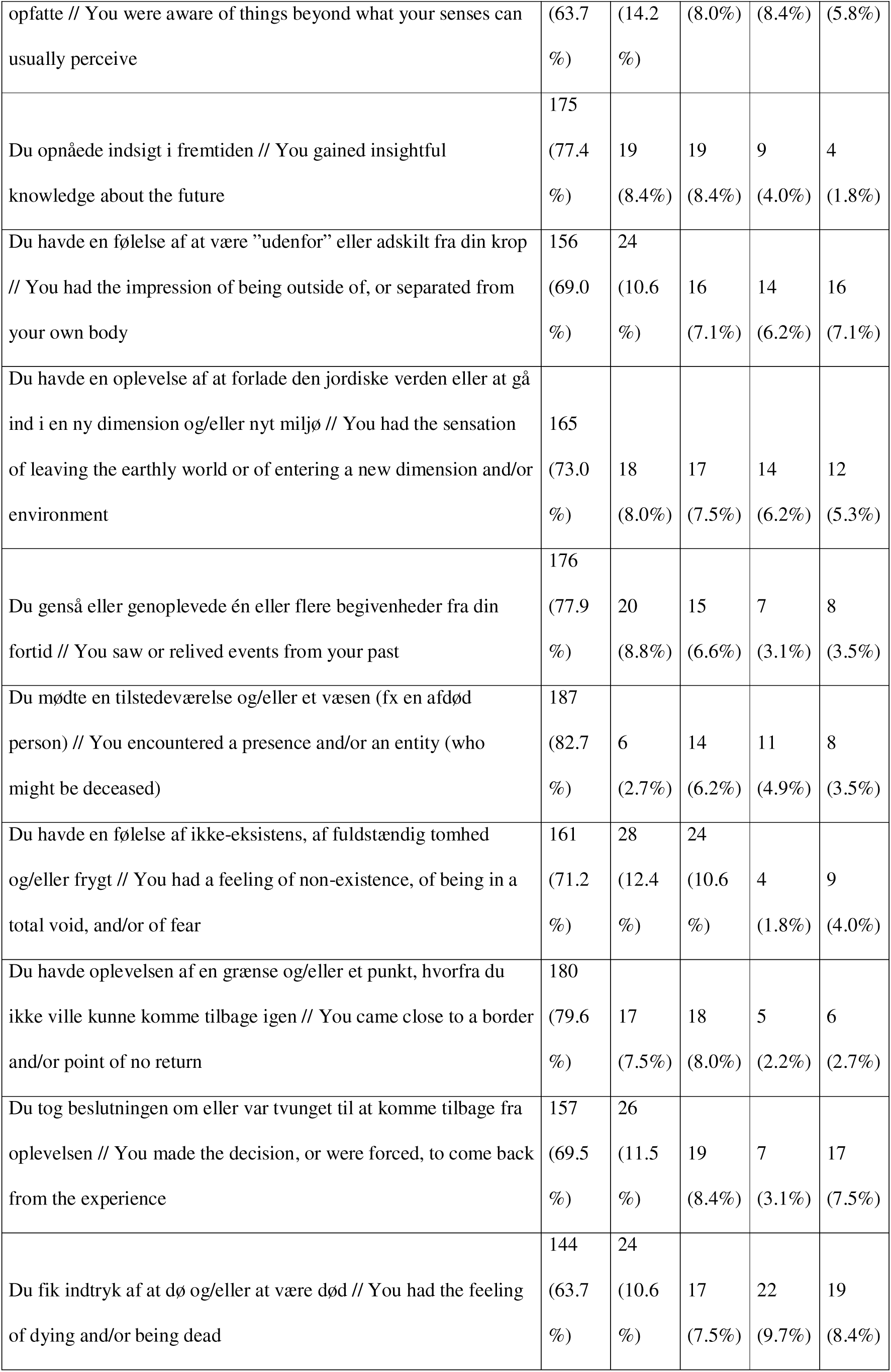
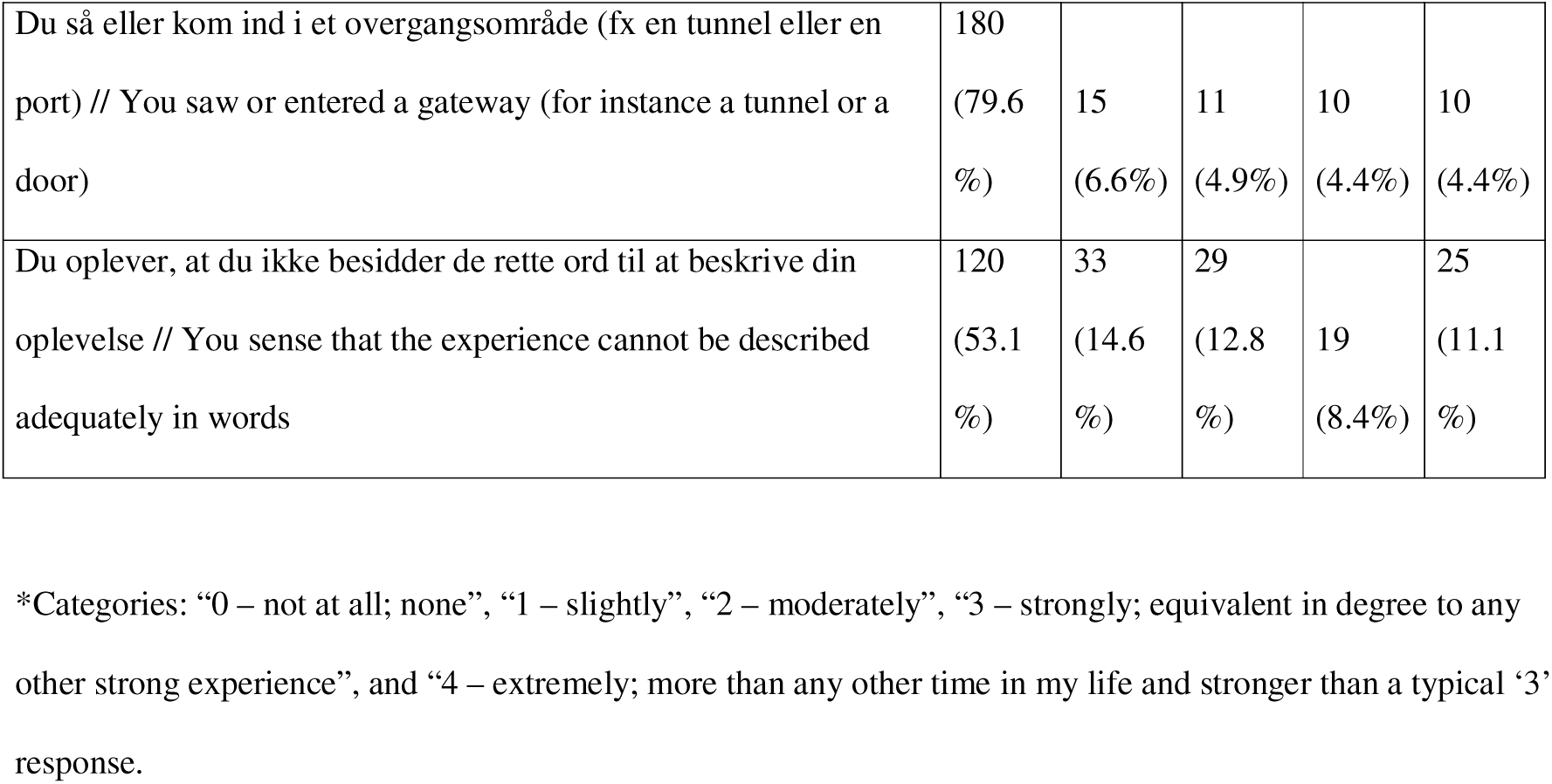
The Danish version of the Near-Death Experience Content scale (NDE-C) (English version in *italics*) and the distribution of responses (n=226).

Psychometric analyses (n=226) of the NDE-C estimated an overall Cronbach’s α = 0.92. The CFA indicated that a five-factor model was superior to a four-factor model in terms of goodness-of-fit, although the overall model fit for both solutions was mixed. The global fit indices were generally poorer than the predefined levels for a good fit, with the CFI at 0.858 and the TLI at 0.831, both falling below their respective thresholds. Furthermore, the RMSEA of 0.093 suggests the model is not a close fit to the data. Conversely, the SRMR of 0.062 is well within the acceptable range, indicating that the model successfully reproduces the observed correlation matrix. Locally, the model exhibits excellent convergent validity, as demonstrated by the overwhelmingly strong factor loadings between observed and latent factors throughout (Figure 1). For instance, item nde2 (“Your thoughts speeded up”) loads at 0.97 on the *Beyond the usual* latent factor, indicating that it is an exceptional measure of its construct. Discriminant validity is also satisfactory with in-between factor correlations of 0.41-0.73, the largest found between *Border* and *Gateway*. While the relationship between these factors is strong, it allows the two factors to remain conceptually distinct and statistically distinguishable.

**Figure 1.**
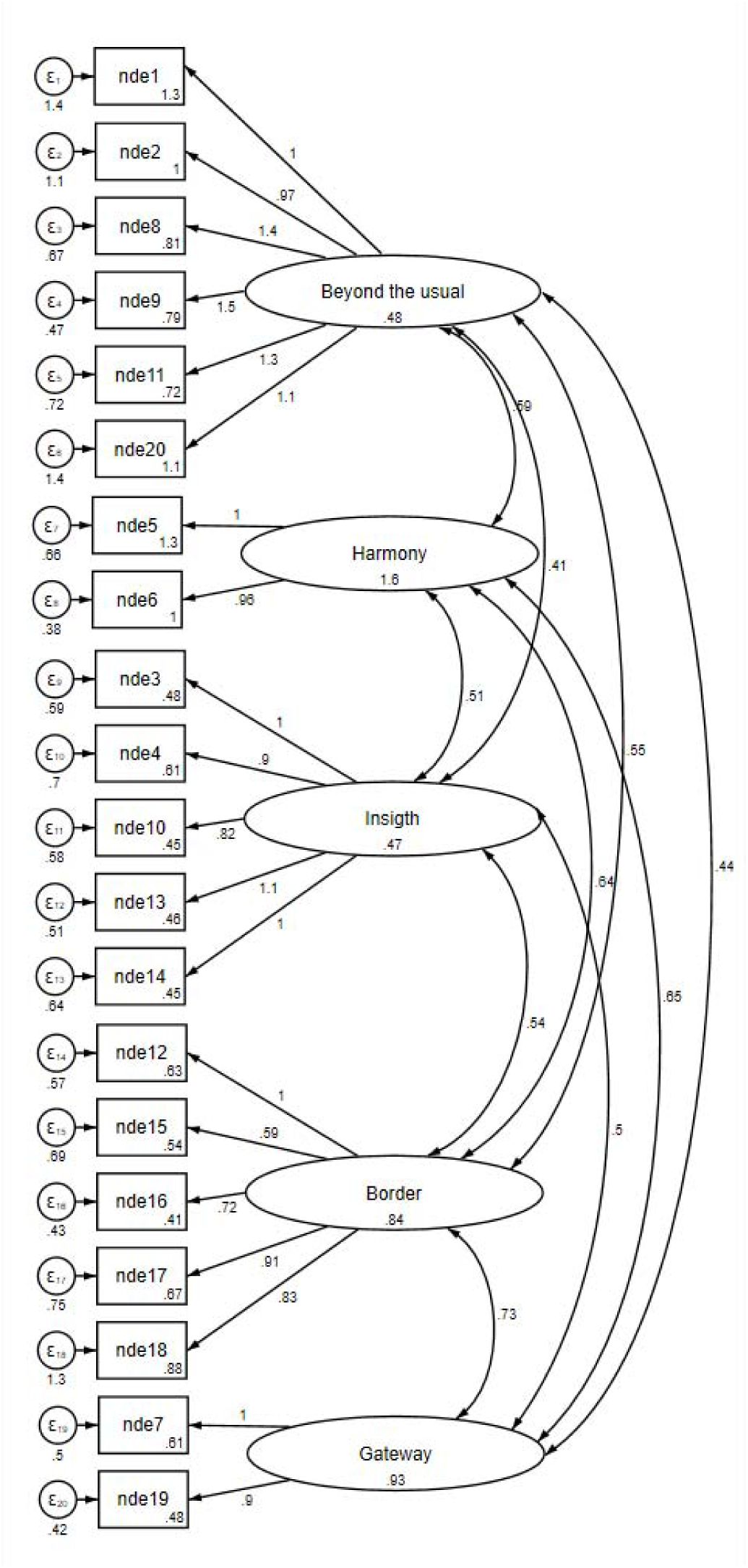
Confirmatory factor analysis model including factor loadings, factor variances, factor covariances, and measurement error variances of the Danish Near-Death Experience Content scale (NDE-C) (n=226).

## DISCUSSION

The initial psychometric analyses of the Danish NDE-C scale suggest that the instrument is a promising tool for studying near-death experiences in Denmark. The scale demonstrated excellent internal consistency but with a mixed structural validity indicated by goodness-of-fit estimates with varying ability to reach a predefined level for a good fit. More research is needed to confirm the dimensionality and generalizability of the Danish NDE-C. Theoretical concerns and ambiguities are discussed.

### Interpretation

The psychometric evaluation of the Danish NDE-C scale indicates that it may be a valid tool for measuring the phenomenology of NDEs among Danish OHCA survivors, albeit with some important caveats. The overall Cronbach’s α of 0.92 indicates excellent internal consistency, slightly higher than that reported in the original development study. This suggests that the items coherently capture a common underlying construct related to NDE content. However, this should also be interpreted with caution, as the scale is multidimensional.

The CFA indicated that a five-factor model provided a better representation of the data than a four-factor model, aligning with the original conceptualization of the scale. However, the global goodness-of-fit indices were mixed. While the SRMR fell within the acceptable range, the CFI, TLI, and RMSEA did not meet conventional thresholds for a close fit. A likely explanation is the relatively low number of respondents (n=226) in relation to the complexity of the model and the number of parameters, which can adversely affect fit indices and model stability. In that light, the model fit could be acceptable, although preliminary, supporting the overall structure while underlining the need for replication in larger samples. In addition, prior qualitative interviews with NDErs using the Danish version of the items indicated satisfactory content validity and acceptability in a secular cultural context ^16^. The observed reduction in model fit compared to the original validation may also stem from differences in sampling strategy. Unlike the current study, the original NDE-C development relied on a specific cohort of individuals who explicitly self-identified as having experienced an NDE.

### Near-death experiences and recalled experiences of death

Reaching a definitive classification consensus on NDEs appears to remain a challenge despite decades of research and efforts to provide a guideline ^4,10^. Thus, recent academic discussion has highlighted and perhaps re-awakened (if ever dormant) conceptual and terminological challenges associated with the term “near-death experience” and proposed to use instead the term “recalled experiences of death” (RED) as a more appropriate umbrella term not risking contamination of experiences with and without real physiological crises ^4^. A central debate concerns whether the phenomenon intrinsically requires a physiological crisis (such as cardiac arrest) – which was the recommendation from Parnia et al. for classifying an experience as an “authentic NDE” or RED ^4^. However, since experiences with similar phenomenology (but not necessarily aftereffects), often termed “NDE-like” experiences (or in some cases spiritually transformative experiences (STEs)), can be induced by non-life-threatening events such as prayer, meditation, or recreational drug use (particularly N,N-Dimethyltryptamine or ketamine), it is questionable to what extent a physiological crisis is a necessary condition ^4,13,20^. While it might be reasonable for methodological reasons to only refer to NDEs if there has been a real physiological crisis (i.e. not to mix what seems to be apples and pears), it might hamper the study of the possible manifestations of human consciousness to insist that authentic NDEs/RED are ontologically different from NDE-like and similar spiritually transformative experiences. Indeed, when tested the NDE-C failed to differentiate between NDE and NDE-like experiences ^13^. Ultimately, it seems various physiological states (i.e. crisis or not) may correspond to various phenomenological states (i.e. NDE/RED/NDE-like or not) and clearly classifying the matrix is complex. The ambiguity in terminology reflects the complexity of the matter.

From this perspective, the Danish NDE-C scale can be viewed as a tool for systematically capturing the content of REDs among OHCA survivors if paired with either qualitative or clinical data providing insight into the context so that a plausible physiological crisis may be confirmed. The scale’s ability to reflect a wide range of cognitive, emotional, perceptual, existential, and spiritual phenomena—including both harmonizing and distressing elements—positions it well within this emerging conceptual framework. Future studies may use the NDE-C in combination with RED-oriented criteria to explore how specific content dimensions relate to clinical variables, neurological status, or long-term psychological and existential outcomes.

A theoretical concern regarding the NDE-C is whether negative experience content reflect the ‘NDE proper,’ or if it stems from misinterpretations of medical interventions and/or emergence from coma ^4^. If the latter is the case, having an item on negative experience content contributing to the overall NDE score would negatively influence the content validity and the cut-off criteria. In a study of distressing NDEs, negative content was woven into the overall NDE “narrative arc” ^8^. Does this reflect various rises and falls in conscious awareness of the physical body rather than the core NDE exhibiting distressing qualia? Whatever the answer, it would always seem wise to probe for negative content during the experience for care purposes. Additionally, without any qualitative supporting data, it is impossible to judge the valence of responding positively to item 15 – fear/void/non-existence, as it is triple-barreled and at least void and non-existence would not necessarily carry a ‘negative’ valence. As the field develops, future research and a better definitional consensus should elucidate these nuances if possible.

### Perspectives

Having a validated Danish measure of NDE content is a key step toward systematically investigating these experiences in a Danish context. NDEs and REDs occupy an important but underexplored intersection between clinical medicine, neuroscience, psychology, and existential or spiritual care. In Denmark, as in many other countries, the field has received limited research attention and minimal dedicated funding, despite growing evidence that NDEs can profoundly affect individuals’ beliefs, values, and mental health.

First, it enables more robust prevalence and phenomenological studies of NDEs/REDs in Danish patient populations, particularly among OHCA survivors. Second, it facilitates cross-cultural comparisons with existing international data, contributing to ongoing debates about the universality versus cultural variability of NDE phenomenology. Third, the tool may inform clinical practice by helping clinicians identify patients who have had such experiences and may benefit from tailored psychological, existential, or spiritual support – regardless of whether the experience was distressing or not. In the long term, systematic use of validated instruments could help integrate NDE- and RED-related care into broader patient-centered and existential care frameworks in cardiology, intensive care, and rehabilitation settings ^21^.

### Strengths and limitations

This study has several strengths and limitations. A key strength is that the sampling frame comprised the full population of OHCA survivors in Denmark from 2016–2020, identified through a nationwide cardiac arrest register rendering sampling bias unlikely. The number of respondents who completed the NDE-C (n=226) is relatively small for a complex CFA model, which may have contributed to the marginal global fit indices and limited the precision of parameter estimates. The present findings should therefore be considered exploratory, and replication in larger samples is warranted before drawing firm conclusions about the factor structure. Recall bias is an inherent concern in retrospective observational studies. However, prior research has shown that memories of NDEs tend to be unusually vivid and stable even with longer recall intervals ^22^. Ideally, the NDE-C would have been validated against the Greyson NDE scale and qualitative narratives. However, this was unfeasible due to space constraints; the scale was embedded within a broader survey prioritizing other primary outcomes. Finally, the sample was restricted to OHCA survivors; thus, the applicability of the Danish NDE-C to populations (e.g. in-hospital cardiac arrest, severe trauma, critical illness) remains to be tested.

## Conclusion

The psychometric evaluation of the Danish Near-Death Experience Content (NDE-C) scale in a sample of out-of-hospital cardiac arrest survivors indicates that the instrument has excellent internal consistency and an acceptable, though not yet good and still preliminary, factor structure aligned with the original validation. The content validity seemed good, although theoretical questions about negative NDE content and the necessity of a physiological crisis or not in defining the phenomenon remains to be further explored with potential implications for the NDE-C. These findings provide an initial foundation for using the NDE-C in research and clinical contexts to systematically assess the phenomenology of near-death experiences, recalled experiences of death, and perhaps other spiritually transformative experiences. However, further studies with larger samples are needed to refine the factor structure, confirm measurement invariance across settings, and explore how specific content dimensions relate to clinical outcomes and existential or spiritual care needs.

## Data Availability

The data are not freely available due to legal and ethical requirements.

## Acknowledgement

The authors would like to thank the study participants who willingly contributed their stories and data to enhance care for cardiac arrest survivors and science at large. Additionally, the authors thank Dr. Sonja Wehberg, University of Southern Denmark, for statistical support and help in designing the study, and Prof. Jens Søndergaard, University of Southern Denmark, for help in designing the study.

## Author contributions

TAS: first draft, design, idea, data collection, data curation, statistical analysis, revision. NCH: design, idea, data collection, revision. All authors accepted the final version.

## Data sharing statement

The data are not freely available due to legal and ethical requirements.

## Generative AI

Large language models were used for rephrasing and adjusting grammar.

## References

1. Holden JM, Greyson B, James D. The handbook of near-death experiences: 30 years of investigation. Praeger Publishers; 2009.

2. Shushan G. Near-Death Experience in Ancient Civilizations: The Origins of the World’s Afterlife Beliefs. Inner Traditions; 2025.

3. Greyson B. A typology of near-death experiences. The American journal of psychiatry. Aug 1985;142(8):967–9. doi:10.1176/ajp.142.8.967

4. Parnia S, Post SG, Lee MT, et al. Guidelines and standards for the study of death and recalled experiences of death--a multidisciplinary consensus statement and proposed future directions. Annals of the New York Academy of Sciences. May 2022;1511(1):5–21. doi:10.1111/nyas.14740

5. Stripp TA. Lights and Landscapes: More[Than[Human Nature in Near[Death Experiences as Reconciliation in a Time of Ecological Crisis. Dialog. 2025;doi:10.1111/dial.12890

6. Kovoor JG, Santhosh S, Stretton B, et al. Near-death experiences after cardiac arrest: a scoping review. Discov Ment Health. May 28 2024;4(1):19. doi:10.1007/s44192-024-00072-7

7. Shushan G. Diversity and similarity of near-death experiences across cultures and history: implications for the survival hypothesis. International review of psychiatry. Feb-Mar 2025;37(2):95–101. doi:10.1080/09540261.2024.2402429

8. Cassol H, Martial C, Annen J, et al. A systematic analysis of distressing near-death experience accounts. Memory. Sep 2019;27(8):1122–1129. doi:10.1080/09658211.2019.1626438

9. Bonenfant RJ. A child’s encounter with the devil: An unusual near-death experience with both blissful and frightening elements. Journal of Near-Death Studies. 1982;20(2):87–100. 10.17514/JNDS-2001-20-2-p87-100.

10. Martial C, Gosseries O, Cassol H, Kondziella D. Studying death and near[death experiences requires neuroscientific expertise. Annals of the New York Academy of Sciences. 2022;1517(1):11–14. doi:10.1111/nyas.14888

11. Holden JM, Kinsey L, Moore TR. Disclosing near-death experiences to professional healthcare providers and nonprofessionals. Spirituality in Clinical Practice. 2014;1(4):278–287. doi:10.1037/scp0000039

12. Greyson B. The near-death experience scale. Construction, reliability, and validity. The Journal of nervous and mental disease. Jun 1983;171(6):369–75. doi:10.1097/00005053-198306000-00007

13. Martial C, Simon J, Puttaert N, et al. The Near-Death Experience Content (NDE-C) scale: Development and psychometric validation. Consciousness and cognition. Nov 2020;86:103049. doi:10.1016/j.concog.2020.103049

14. Buer O, Kalfoss M, Weisaeth L, Bendz B. Investigating near-death experiences. Tidsskrift for den Norske laegeforening : tidsskrift for praktisk medicin, ny raekke. Dec 2016;136(23-24):1968–1969. Kartlegging av naer doden-opplevelser. doi:10.4045/tidsskr.16.0953

15. Stripp TK, Wehberg S, Büssing A, et al. Protocol for EXICODE: the EXIstential health COhort DEnmark—a register and survey study of adult Danes. BMJ Open. 2022;12(6):e058257. doi:10.1136/bmjopen-2021-058257

16. Stripp TA, Viftrup DT, Nissen RD, Wehberg S, Sondergaard J, Hvidt NC. Testing the acceptability and comprehensibility of a questionnaire on existential and spiritual constructs in a secular culture through cognitive interviews. Survey Research Methods. 2023;17(1):75–89. doi:10.18148/srm/2023.v17i1.7971

17. Pedersen CB. The Danish Civil Registration System. Scand J Public Health. Jul 2011;39(7 Suppl):22–5. doi:10.1177/1403494810387965

18. Kline RB. Principles and Practice of Structural Equation Modeling, Second Edition. Guilford Publications; 2005.

19. Hooper D, Coughlan J, Mullen M. Structural Equation Modeling: Guidelines for Determining Model Fit. The Electronic Journal of Business Research Methods. 11/30 2007;6doi:10.21427/D7CF7R

20. Rominger R. Integration of Spiritually Transformative Experiences: Models, Methods, and Research. Journal of Near-Death Studies. 2013;31(3)

21. Stripp TK, Joshi VL. [A review of existential concerns of cardiac arrest survivors and implications for rehabilitation: do we need a new tool in the toolbox?]. Tidsskrift for Forskning i Sygdom og Samfund. 2023;20(38):25–48. doi:10.7146/tfss.v20i38.131489

22. Greyson B. Consistency of near-death experience accounts over two decades: are reports embellished over time? Resuscitation. Jun 2007;73(3):407–11. doi:10.1016/j.resuscitation.2006.10.013

